# Socioeconomic inequality in the double burden of child malnutrition in the Eastern and Southern African Region

**DOI:** 10.1101/2022.03.30.22273164

**Authors:** Rishi Caleyachetty, Niraj Kumar, Hana Bekele, Semira Manaseki-Holland

## Abstract

Socioeconomic inequalities in the double burden of child malnutrition threatens global nutrition targets 2025, especially in Eastern and Southern Africa. We aimed to quantify these inequalities from nationally representative household surveys in 13 Eastern and Southern African countries between 2000 and 2018. 13 of the latest Demographic and Health Surveys including 72,231 children under five year olds were studied. Prevalence of stunting, wasting and overweight (including obesity) were disagregated by wealth quintiles, maternal education categories and urban-rural residence for visual inspection of inequalities, and the slope index of inequality (SII) and the relative index of inequality (RII) were estimated for each country. Country-specific estimates were pooled using random-effects meta-analyses. Regional stunting and wasting prevalence was higher among children living in the poorest households, with mother’s with the lowest educational level and in rural areas. In contrast, regional overweight (including obesity) prevalence was higher among children living in the richest households, with mother’s with the highest educational level and urban areas. Tackling social inequalities in the distribution of the double burden of malnutrition among children in the Eastern and Southern African region will require strategies that address the reasons socially disadvantaged children become more exposed to stunting or wasting.

## Introduction

The African region has the highest burden of childhood stunting and one of the highest burdens of childhood overweight in Africa.^1^ Importantly, the region is off track to achieve the UN Sustainable Development Goal (SDG) 2 aim to end all forms of hunger and malnutrition by 2030.^2^ The double burden of malnutrition, is considered a major global health challenge for African countries,^3^ particularly those in Eastern and Southern Africa. The prevalence of malnutrition is 34.5% in Eastern Africa for stunting and 10.2% in Southern Africa for overweight (including obesity), compared to 29.1% for stunting and 4.7% for overweight (including obesity) in Africa.^1^

Following on from the 2008 WHO Commission on Social Determinants of Health and 2011 Rio Political Declaration on Social Determinants of Health, another important goal for the African region is tackling social inequalities in health.^4^ Considering that the double burden of malnutrition is inextricably bound to socioeconomic conditions,^5^ monitoring and description of child malnutrition inequalities will be important to serve evidence-based program and policy decisions in the Eastern and Southern African region. Several measures of social inequality in health reflecting its different dimensions have been described.^6^ In the context of child malnutrition, absolute inequality measures reflects the magnitude of difference in malnutrition prevalence between two subgroups and relative inequality measures show proportional differences in child malnutrition prevalence among subgroups. The choice of measure is not value-neutral and influences our understanding of which populations may be actually experiencing a higher malnutrition burden. Therefore, both relative and absolute measures to monitor social inequalities are recommended.^7^

A recent study examined global inequalities in household-level double burden of malnutrition and found the probability of double burden of malnutrition higher among richer households in poorer low-income and middle-income countries.^8^ Their definition of double burden of malnutrition was expressed as a stunted child with an overweight mother. The objective of reducing child malnutrition may not necessarily be compatible with the objective of reducing socioeconomic inequalities in child malnutrition. For example, Africa has observed spectacular gains in reducing stunting, however these gains may have not benefited every child equally.^9^ In the Eastern and Southern African region, there has not been to date a comprehensive large-scale assessment of socioeconomic inequalities in the population-level double burden of child malnutrition. Such an assessment would be valuable to support countries moving towards regional nutrition equity.

We use nationally representative household surveys across 13 Eastern and Southern African countries between 2003-2018 to quantify socioeconomic inequalities in the double burden of malnutrition among children under five years old.

## Methods

We searched for data from the most recent Demographic and Health Survey (DHS)^10^ within countries in the Eastern and Southern African region with data available on length/height and weight. In all countries, these surveys followed the same standardised procedures. Complete descriptions of country DHS sampling, questionnaire validation, data collection methods, and data validation procedures are published elsewhere.^10^ Our analytical sample included alive children under age five years with valid weight and height measurement, and living with their mother. The datasets used for this project are available at the DHS Program website (https://www.dhsprogram.com/data/available-datasets.cfm).

### Ethics statement

The DHS was approved centrally by ICF International (Calverton, MD, USA) institutional review board and by individual review boards within every participating country. Informed verbal consent was obtained from all participants during surveys and data were released in deidentified form. A parent or guardian provided verbal consent prior to participation by a child.

### Anthropometric measurements

DHS include data about each child’s age (in months and years) and measured height/weight. Trained survey staff weighed and measured the length/height of each child. Height-for age and BMI-for-age z-scores were calculated using the ‘zscore06’ STATA command, which uses WHO 2006 growth standards. Stunting was defined as z scores for height-for-age (HAZ) of <-2SD. Wasting was defined as z scores for weight-for-height (WHZ) <-2 SD. Overweight was defined as z scores for WHZ >2 SD and ≤ 3 SD of the median and obese was defined as z scores for WHZ > 3 SD.

### Socioeconomic characteristics

The economic status of households was based on asset indices. Household questionnaires collect information on household assets (such as televisions, refrigerators, and other appliances), dwelling characteristics (materials used for the walls, floor and roof, presence of electricity, water supply and sanitary facilities) and other variables associated with wealth (such as ownership of the house, of land or livestock). A wealth index is constructed using household asset data via principle component analysis, which can be split into quintiles, where the first quintile (Q1) represents the approximately 20% poorest households in the survey sample and the fifth quintile (Q5) represents the richest. Maternal education was self-reported and categorised in three groups: none (no formal education); primary (any primary education, including completed primary education); and secondary or higher (any secondary education, including complete secondary). Urban or rural location were defined defined by the national census or statistical bureau in each country at the time the survey was conducted.

### Statistical analysis

Regional prevalence estimates and 95% confidence intervals (CIs) for stunting, wasting, overweight (including obesity), concurrent stunting and wasting, and concurrent stunting and overweight (including obesity) in children <5 years were calculated. First, we calculated country-specific malnutrition estimates. The DHS used complex sample designs therefore we accounted for stratification and clustering. Meta-analysis of country-specific estimates were performed to calculate the regional prevalence estimates and 95% CIs. Variances of the raw proportions were stabilized with a double arcsine transformation and then pooled on the basis of a random-effects model.

Social inequalities in the population-level double burden of child malnutrition were first visualized using equiplots. Equiplots allow comparison of absolute social inequality indicating both the prevalence of malnutrition in each group and the distance between groups, which represents the absolute social inequality. We then estimated using absolute and relative differences between poorest and richest households, lowest and highest maternal education and urban and rural residence. Wealth and education inequalities in the double burden of malnutrition were measured for each country by regression-based inequality measures, the relative index of inequality (RII) and the slope index of inequality (SII), respectively.^11^ A logistic regression model was used to assess the association between malnutrition and wealth or maternal education and to generate the SII and RII values and 95% CIs. Social inequalities on both scales were reported because conclusions can be skewed when only one or the other is used.^12^ Both these measures take into account the size of the population across wealth and education groups. The SII can be interpreted as the defined as the absolute difference in the malnutrition prevalence between the poorest and richest or mothers with the lowest education and mothers with the highest education level (across the entire socioeconomic or maternal education distribution). SII can range from -1 to 1. If there is no inequality, SII takes the value zero. Positive SII values indicate that the child malnutrition indicator is concentrated in the most socially advantaged group (i.e. richest or mothers with the highest education). The RII is defined as the ratio of the estimated prevalence of malnutrition between the poorest and the richest or mothers with the lowest education and mothers with the highest education (across the entire socioeconomic or maternal education distribution). If there is no inequality, RII takes the value one. An RII value great than 1 indicates the child malnutrition indicator is concentrated among the most socially advantaged group. We used random-effects meta-analysis to average inequality estimates across countries in the Eastern and Southern African region. All of the statistical analyses were performed with the use of STATA version 16.1 (StataCorp). We used the svy command to account for complex survey sampling designs and the sampling weights for all countries’ surveys.

## Results

Demographic Health Survey datasets from 2003-2018 were available for 17 Eastern and Southern African countries. 15 out of 17 had available anthropometric data however access to two DHS datasets were restricted. In total, 13 out of 17 (77.0%) countries and 116,747 children living with their mother were eligible for inclusion in our study. Of the children aged 0-5 years and living with their mother (n=75,894), 4.0% (n=3037) with implausible HAZ and 0.8% (n=626) implausible WHZ. 72,231 children (95.2%) were included in our final analytical sample.

Table 1 shows the country-level characteristics of participants. The mean age of children was 28.4 months (range: 26.0-29.8 months). The proportion of children with mothers who had no formal education ranged from 1.0% (Lesotho) to 48.0% (Comoros), and the proportion of children who lived households in the lowest household wealth quintile ranged from 21.6% (Lesotho) to 25.8% (Comoros). The proportion of children living in urban areas ranged from 13.0% (Malawi) to 56.9% (South Africa).

**Table 1.** Characteristics of most recent Demographic Health Surveys in Eastern and Southern African region

| Country | Year | Children < 5 years (n) | Analytic sample (n) | Response Rate (%) | Age (months) | Living in urban area (%) | Lowest household wealth quintile (%) | No maternal education (%) |
| --- | --- | --- | --- | --- | --- | --- | --- | --- |
| Comoros | 2012 | 2,882 | 2,432 | 84.4 | 28.6 | 26.6 | 25.8 | 48.0 |
| Eswatini | 2006 | 2,220 | 2,042 | 92.0 | 27.7 | 18.7 | 22.0 | 9.2 |
| Kenya | 2015 | 19,255 | 18,648 | 96.8 | 29.1 | 34.3 | 24.2 | 11.9 |
| Lesotho | 2014 | 1,367 | 1,303 | 95.3 | 27.0 | 27.3 | 21.6 | 1.0 |
| Malawi | 2015 | 5,357 | 5,116 | 95.5 | 29.6 | 13.1 | 23.8 | 13.0 |
| Mozambique | 2011 | 9,772 | 9,363 | 95.8 | 27.8 | 26.9 | 23.8 | 37.7 |
| Namibia | 2013 | 1,945 | 1,800 | 92.5 | 26.0 | 42.6 | 25.2 | 6.9 |
| Rwanda | 2014 | 3,615 | 3,544 | 98.0 | 28.4 | 16.5 | 24.8 | 14.3 |
| South Africa | 2016 | 1,451 | 1,070 | 73.7 | 29.8 | 56.9 | 24.1 | 19.8 |
| Tanzania | 2015 | 9,196 | 8,940 | 97.2 | 28.0 | 25.9 | 24.7 | 21.4 |
| Uganda | 2016 | 4,506 | 4,382 | 97.2 | 28.8 | 20.4 | 22.7 | 11.1 |
| Zambia | 2018 | 9,094 | 8,694 | 95.6 | 28.8 | 35.0 | 25.1 | 10.1 |
| Zimbabwe | 2015 | 5,234 | 4,897 | 93.6 | 29.4 | 29.6 | 24.4 | 1.2 |

Fig 1 shows the regional prevalence estimates for child malnutrition. Overall, stunting prevalence was 30.5% (95% CI 27.7,33.4), with prevalence ranging from 22.3% (Namibia) to 39.3% (Mozambique) (S1 Table); wasting prevalence was 4.6% (95% CI: 3.7-5.6), with prevalence ranging from 2.3% (Rwanda) to 12.3% (Comoros) (S2 Table); and overweight (including obesity) prevalence was 6.5% (95% CI 5.2,7.9), with prevalence ranging from 3.6% (Tanzania) to 12.9% (South Africa) (S3 Table). The pooled prevalence of concurrent stunting and wasting was 1.1% (95% CI 0.8,1.4), with prevalence ranging from 0.4% (South Africa) to 2.1% (Comoros) (S4 Table). The pooled prevalence of concurrent stunting and overweight (including obesity) was 2.6% (95% CI 1.9, 3.5), with the prevalence varying across countries, ranging from 1.1% (Kenya) to 5.1% (Comoros) (S5 Table).

**Fig 1.**
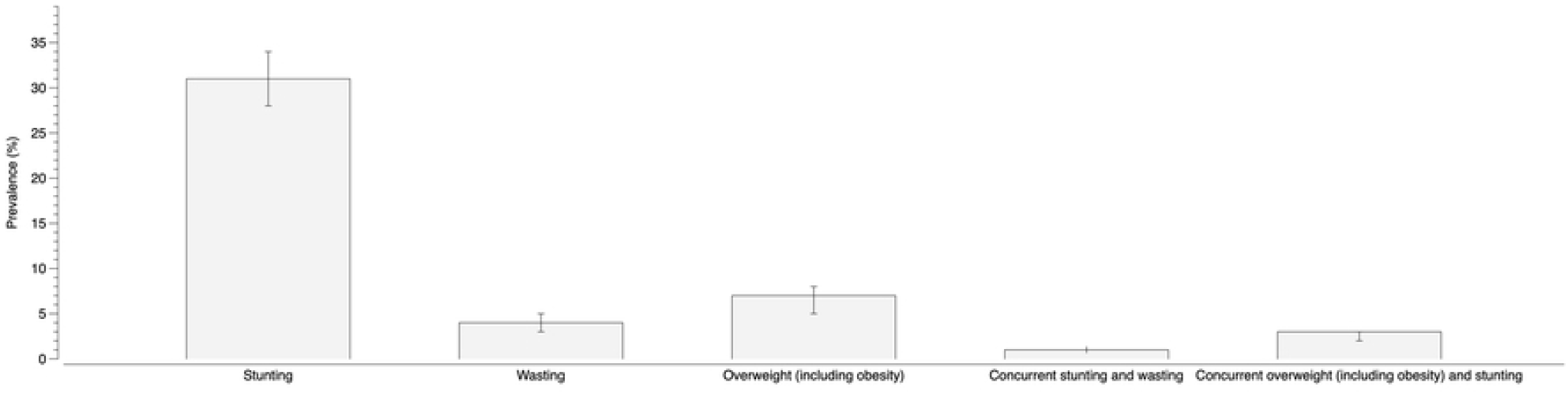
Pooled Eastern and Southern African region prevalence es.

Fig 2 shows the prevalence of adolescent overweight (including obesity) by stunting or wasting prevalence, with each country in the region represented by one square. The vertical and horizontal lines separate four quadrants. The plot identifies 23% (3 of 13) of Eastern and Southern African countries had prevalences of overweight (including obesity) and stunting or wasting greater than the overall pooled prevalence estimates for overweight (including obesity) and stunting or wasting, respectively.

**Fig 2.**
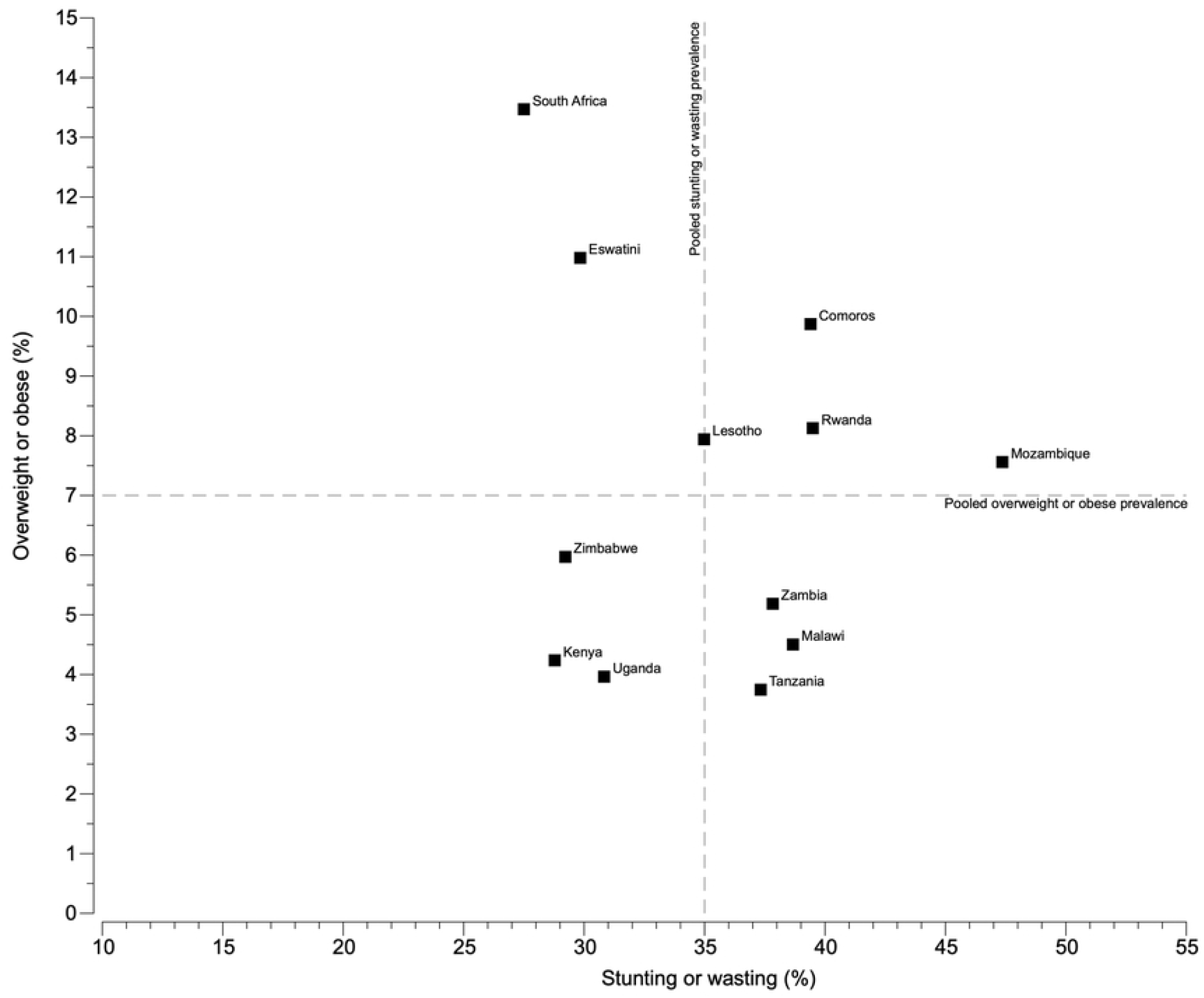
Scatterplot of overweight or obesity prevalence by stunting.

Fig 3 presents equiplots of regional stunting prevalence by wealth quintiles, categories of formal maternal education, and urban-rural residence. Higher stunting prevalence was found among children belonging to the poorest households, mother’s with the lowest educational level and rural residence. Lesotho represents an extreme example with the widest gaps in stunting prevalence by wealth (30.5% points; p<0.001) and maternal education (54.6% points; p<0.001) (S6-S7 Tables). Malawi showed the widest gap in stunting prevalence by urban-rural residence gap (13.7% points; p<0.001)(S8 Table).

**Fig 3.**
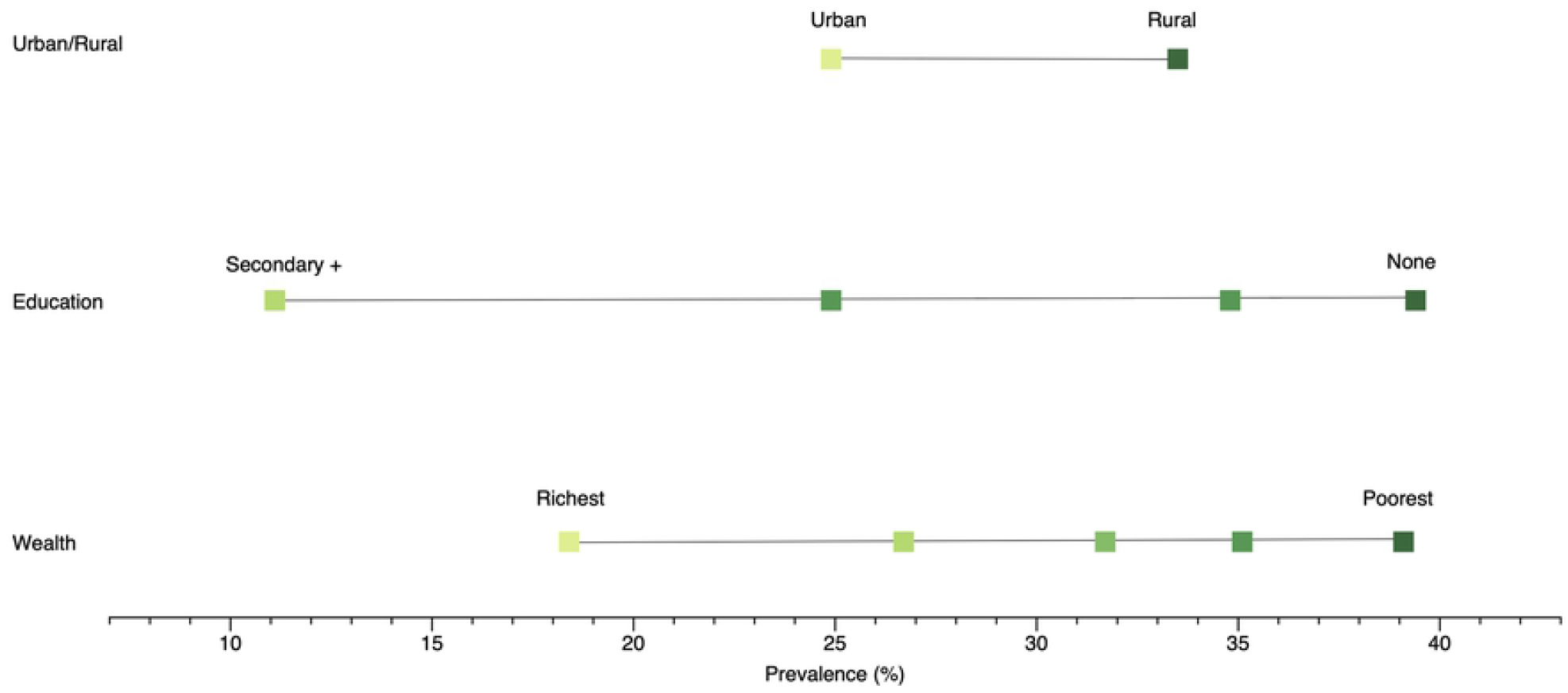
Equiplot showing stunting prevalence by wealth quintiles.

Fig 4 presents equiplots of regional wasting prevalence by wealth quintiles, categories of formal maternal education categories, and urban-rural residence for the region. Higher wasting prevalence was found among children belonging to the poorest households, mother’s with the lowest educational level, and rural residence. Mozambique and Namibia represents extreme examples with the widest gaps in wealth (7.1% points; p<0.001) and maternal education gaps (16.2% points; p< 0.001) (S9-S10 Tables). Mozambique showed the widest gap in wasting prevalence by urban-rural residence (3.0%; p<0.001) (S11 Table).

**Fig 4.**
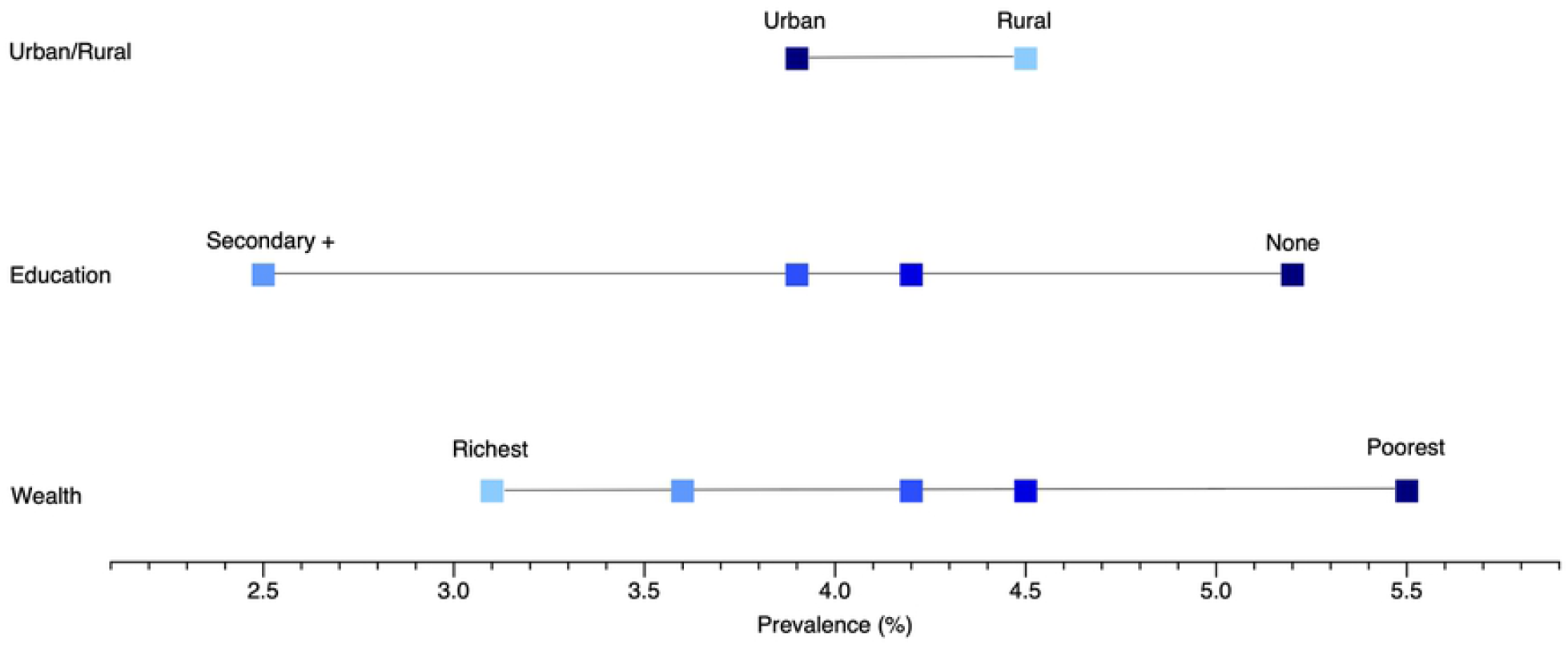
Equiplot showing wasting prevalence by wealth quintiles.

Fig 5 presents equiplots of overweight (including obesity) prevalence by wealth quintiles, categories of formal maternal education categories, and urban-rural residence for the region. Higher overweight (including obesity) prevalence was found among children belonging to the richest households, mother’s with the highest educational level and urban residence. Eswatini represents an extreme example, with the the widest gaps in overweight (including obesity) prevalence by wealth (6.1% points; p=0.058), maternal education (12.2% points; p<0.001) and urban-rural residence (6.8% points; p=0.002) (S12-14 Tables).

**Fig 5.**
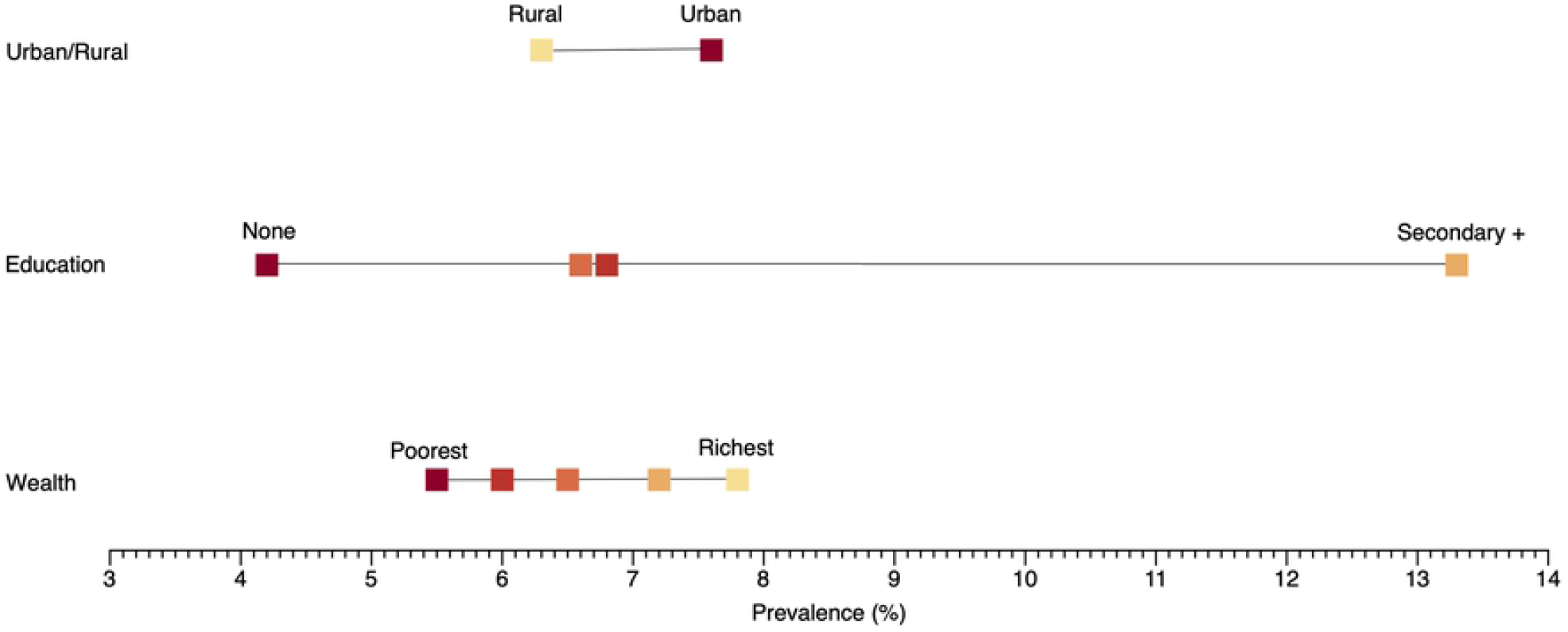
Equiplot showing overweight (including obesity) prevalence.

Fig 6-9 show the mean values of stunting, wasting and overweight (including obesity) prevalence respectively, and summary inequality measures (that take the whole socioeconomic distribution into account). Significant levels of relative and absolute inequalities in favour of the poorest households and least educated mothers were observed for child stunting and wasting. In the absolute scale, the largest wealth inequality in stunting prevalence was in Rwanda (SII= -0.36; 95% CI 0.41,-0.31) and in the relative scale, the largest wealth inequality was in Namibia (RII= 0.36, 95% CI 0.25, 0.47). The largest maternal education inequality in stunting prevalence was in Rwanda on the absolute scale (SII=-0.33, 95% CI -0.39, -0.26) and in relative scale Namibia on the (RII=0.33, 95% CI 0.22, 0.43). Kenya had the largest inequalities in child wasting prevalence by household wealth quintile (SII: -0.10, 95% CI -0.12, -0.09; RII: 0.17, 95% CI 0.13, 0.21) and maternal education (SII: -0.13, 95% CI -0.15, -0.11; RII: 0.10 95% CI 0.08, 0.13). In contrast, there were inequalities in the other direction for overweight (including obesity), that is, overweight (including obesity) was concentrated towards the richest wealth quintiles children and higher maternal education. In the absolute scale, the largest wealth inequality in overweight (including obese) prevalence was in Comoros (SII: 0.09, 95% CI 0.04, 0.13) and in the relative scale, Kenya (RII: 4.46, 95% CI 3.24, 4.67). The largest maternal education inequality in overweight (including obese) prevalence on the absolute scale was in Eswatini (SII: 0.11, 95% CI 0.05, 0.16) and Kenya on the relative scale (RII: 4.35, 95% CI 3.09, 5.60).

**Fig 6.**
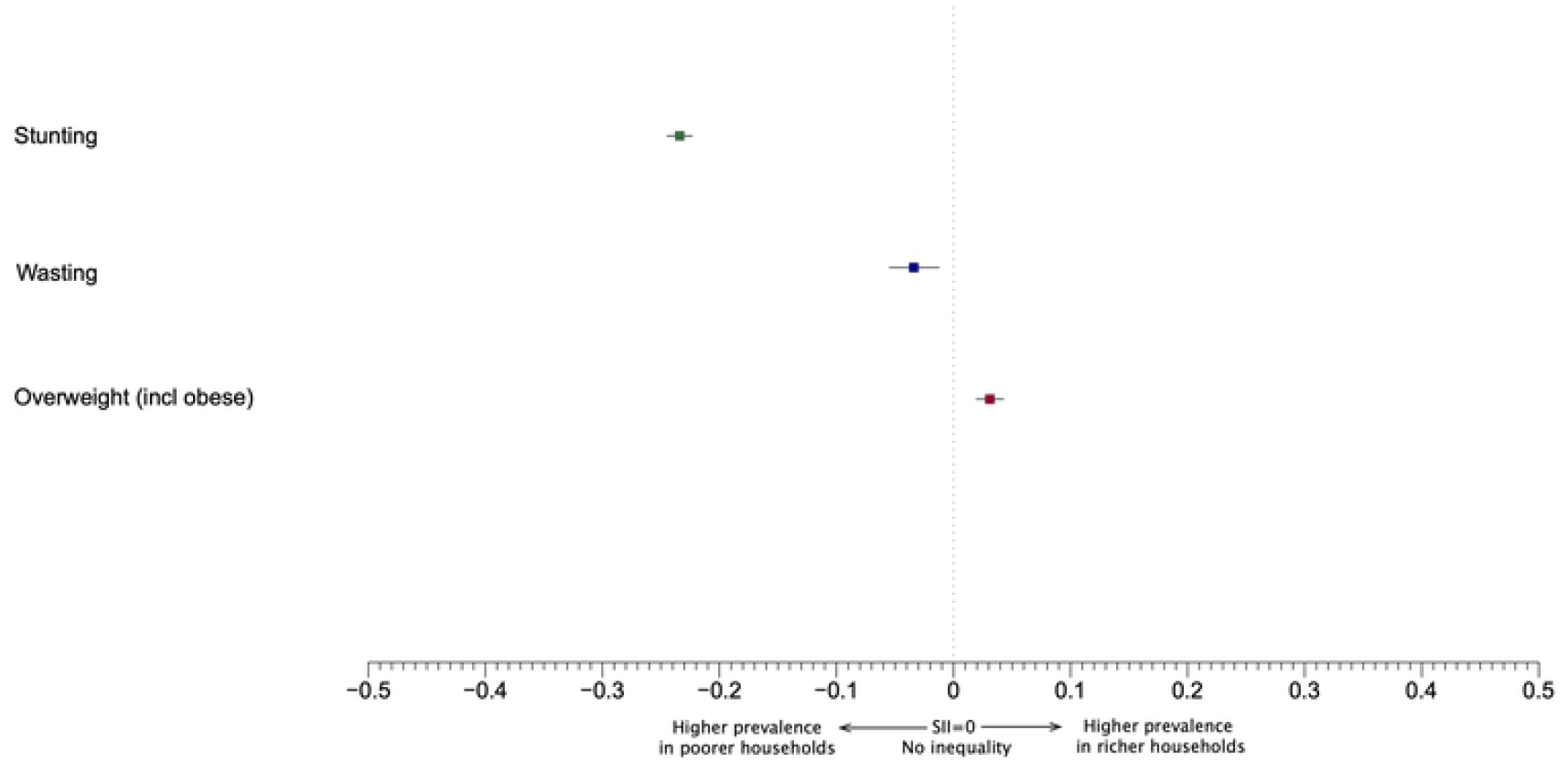
Absolute (Sil) inequalities in the double burden of malnutrition.

**Fig 7.**
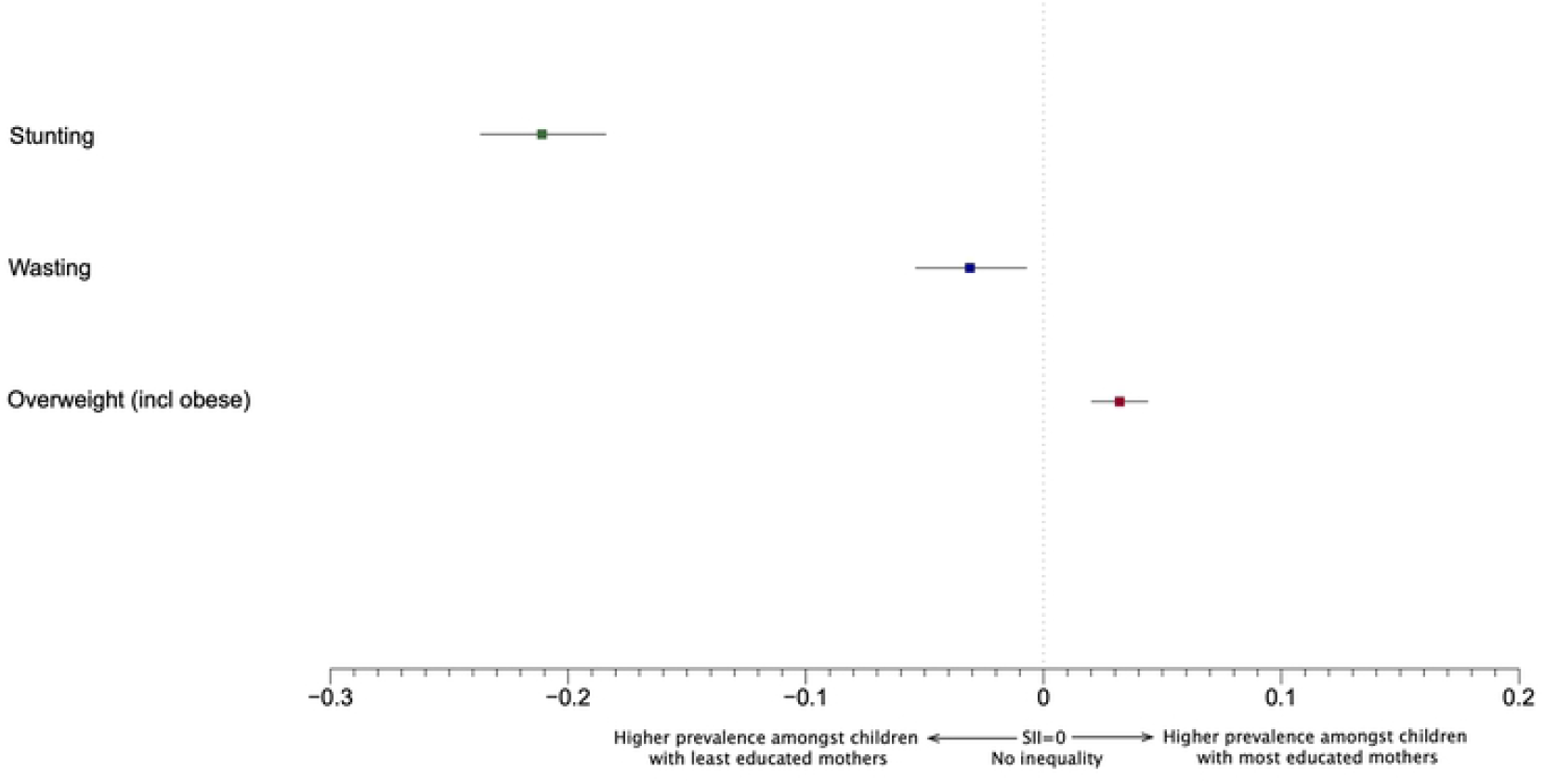
Absolute (Sil) inequalities in the double burden of malnutrition.

**Fig 8.**
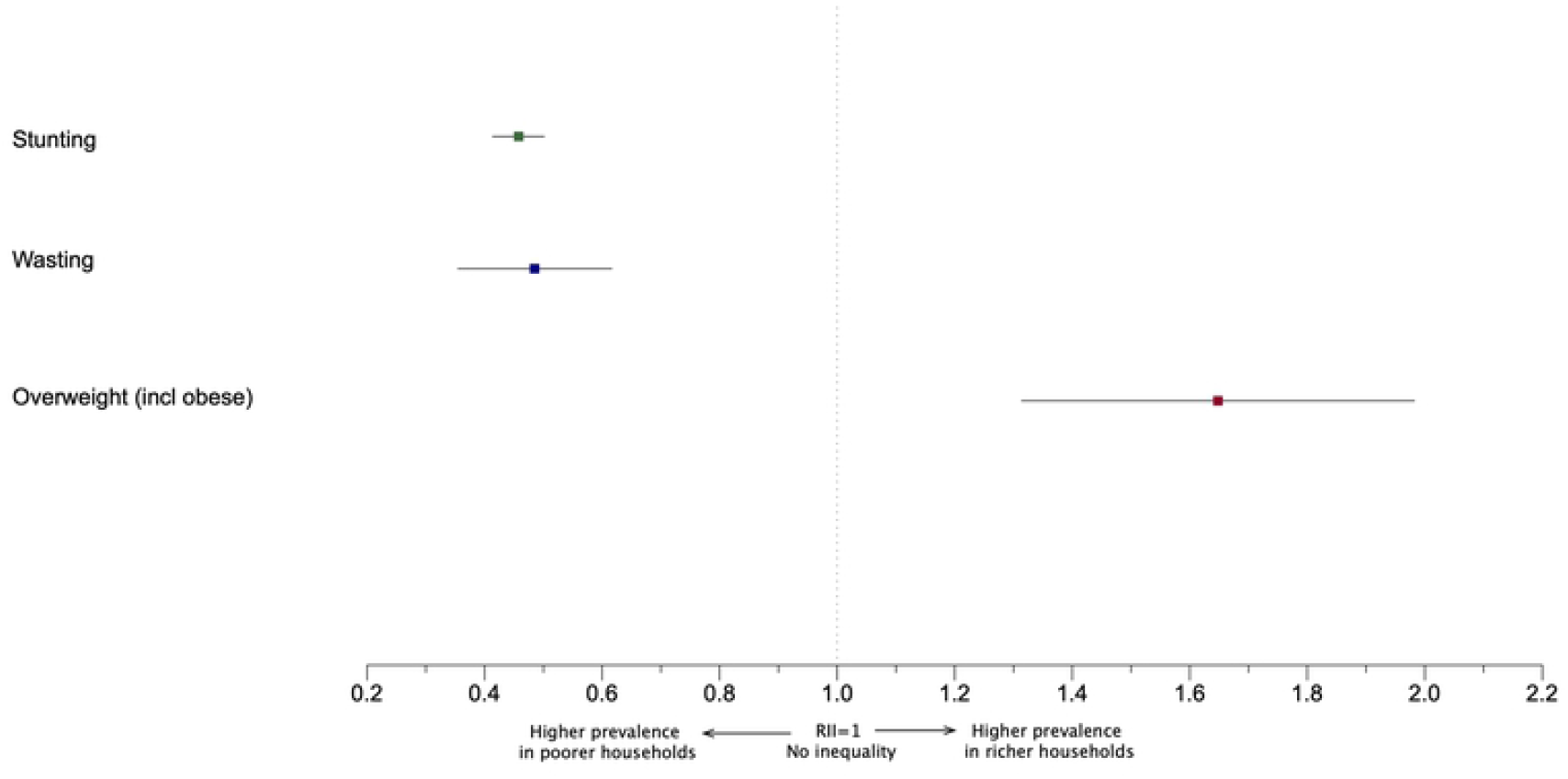
Relative (RII) inequalities in the double burden of malnutrition.

**Fig 9.**
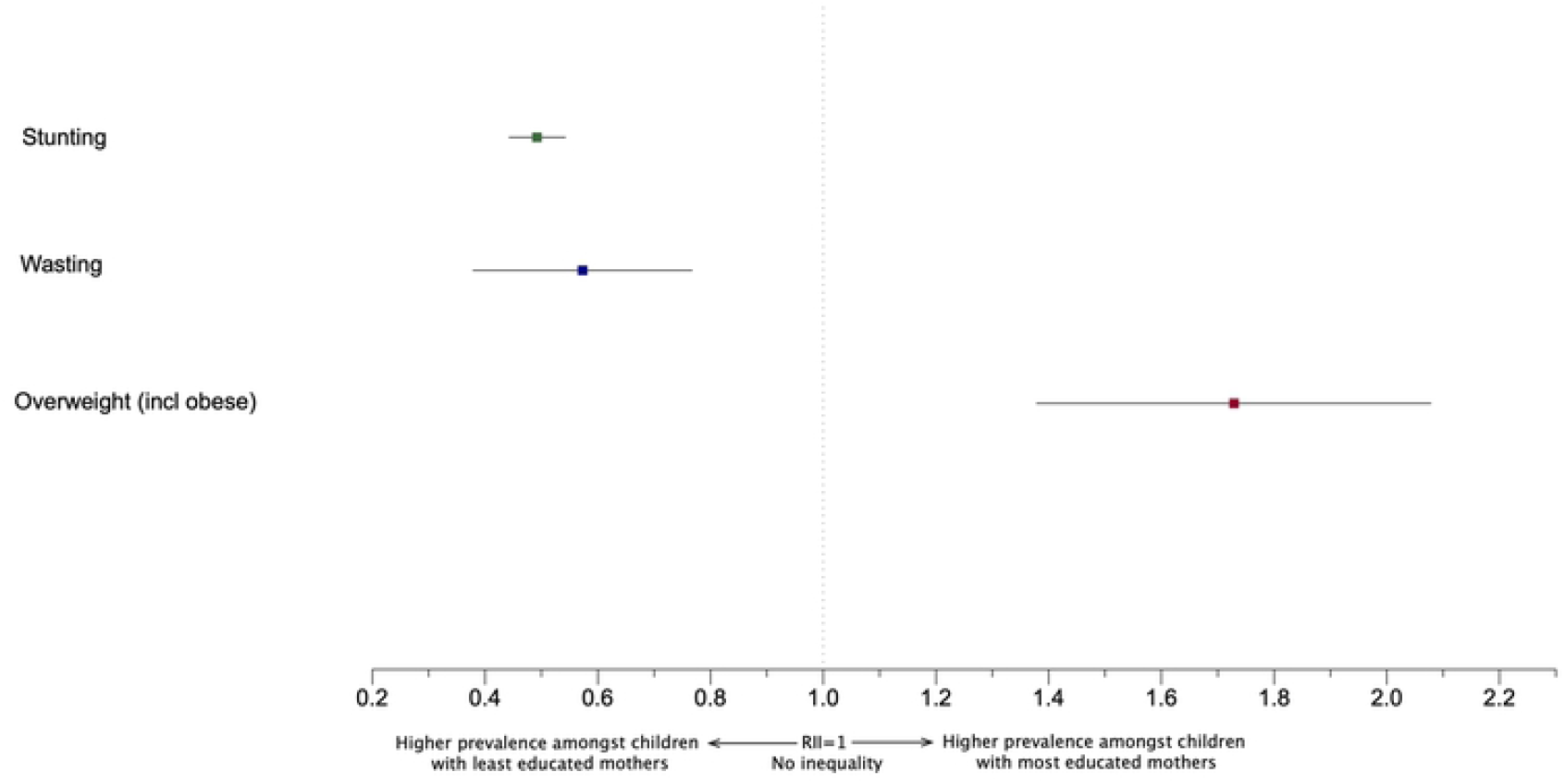
Relative (RII) inequalities in the double burden of malnutrition.

## Discussion

To our knowledge, this study is the first to systematically identify the magnitude of social inequalities in the double burden of malnutrition among children under five years across the Eastern and Southern African region. We documented important levels of socioeconomic inequalities both on the relative and absolute scales in stunting, wasting and overweight (including obesity), as well as urban-rural inequalities. The magnitude of these inequalities varied across countries and malnutrition indicators. Regionally, stunting and wasting prevalence were concentrated in the lowest wealth quintile and maternal education level, and rural areas. These inequalities were in the other direction for overweight (including obesity).

A study of 55 low-income and middle-income countries (LMICs) between 1992-2018 examining inequalities in double burden of malnutrition found a higher probability of the double burden of malnutrition among richer households in poorer LMICs and poorer households in richer LMICs.^8^ The authors defined the double burden of malnutrition at the household level, expressed as a stunted child with an overweight mother. However, there is strong evidence that stunted child/overweight mother pairs represents a statistical artefact and not a distinct entity.^13^ Our study assessed socioeconomic inequalities in the double burden of child malnutrition at the population level, which is a common double burden of malnutrition operational definition.^14^ A systematic review examining socioeconomic status and overweight or obesity among older children (5-19 years) in Sub-Saharan Africa used the RII to assess socioeconomic inequalities. The school-age children from higher socioeconomic households tended to be overweight and obese.^15^ We found similar wealth and education inequalities using both simple and complex absolute and relative measures in the Eastern and Southern African region for children under five year olds. In highly developed countries, a higher prevalence of child overweight and obesity children is typically found in lower socioeconomic groups.^16^ Eastern and Southern African households of lower socioeconomic status, may have financial constraints that prevent purchasing of high-energy density foods with a consequence that the overweight and obesity prevalence levels are kept to a minimum in these households. Additionally, parent’s socio-cultural beliefs may influence their child’s body shape ideals. In some sub-Saharan African countries, overweight and obesity have been historically been considered to be a sign of wealth.^15^

Our study found the Eastern and Southern African region is burdened by multiple forms of malnutrition. The prevalence of stunting among children was 30.4%, the prevalence of wasting was 4.6%, whereas 6.5% were overweight (including obesity). A much smaller proportion of children also had concurrent stunting and wasting (1.1%) and concurrent stunting and overweight (2.6%). The presence of a double burden of malnutrition at a population level, is likely to reflect the region’s ongoing challenges with poverty, food insecurity, infectious diseases, droughts, floods and conflict as well as the presence of the obesogenic environment driven by globalization and rapid urbanization. Households of lower socioeconomic households may not have access to resources (money, knowledge, prestige, power and beneficial connections) that could better able them to avoid undernutrition and therefore unable to avoid child undernutrition.

Several limitations, however, should be considered when interpreting our findings. The Eastern and Southern African region encompasses 24 countries, however our findings are for 13 countries of 17 in the Eastern and Southern African region where DHS is available. A direct indicator of household income is not collected in the DHS and the asset-based household wealth index was used. While this is an imperfect measure financial resources, it is frequently used and judged to be superior to income in lower income countries.^17^ Between country comparisons may be affected by the period of data collection for each national household survey (from 2003 to 2018). Social reform and safety net programmes in the region have increased over this study period and may have reduced inequalities over time.^18^ Despite these limitations, our study has several strengths. Our analysis of socioeconomic inequalities and urban/rural inequalities in child malnutrition in Eastern and Southern African region are based on a relatively large and diverse sample of children from 13 nationally representative household surveys with uniformity of information based on the same questions and procedures in all countries. We used both simple and complex measures to show inequality. Simple measures only compare two subgroups. For example, we used equiplots to compare the richest and poorest. However, simple measures cannot describe how all groups change. We also used complex measures (including SII and RII) which indicated inequality across the whole population between the most and the least disadvantaged.

Addressing social inequalities in the distribution of the double burden of malnutrition among children in the Eastern and Southern African region requires strategies that address the reasons certain subpopulations became more exposed to these nutrition problems, while also avoiding strategies that solve one nutrition problem while worsening another one.^5^ Any attempt to analyse and act on inequalities in the double burden of child malnutrition in the Eastern and Southern African region, however requires the initial acknowledgement that tackling overweight (including obesity) among socially advantaged children, is not part of the “health inequalities” agenda, which focuses on the improving the health of socially disadvantaged.^19^ Most countries also do not have a robust routine data collection systems for child malnutrition,^20^ and therefore cannot adequately and frequently monitor socioeconomic gradients in child malnutrition. Countries may therefore not have programmes and policies that are also tailored to the desired population.

Given the recent coronavirus disease 2019 (COVID-19) pandemic, this is likely to be a time of worsening socioeconomic conditions.^21^ Therefore, it is important that there be an ongoing conversation about measuring and acting on the socioeconomic inequalities of the double burden of child malnutrition in the Eastern and Southern African region, as there much to debate and resolve.

## Data Availability

The datasets used for this project are available at the DHS program website (https://www.dhsprogram.com/).

https://www.dhsprogram.com/data/available-datasets.cfm

